# Fasudil attenuates disease spreading in ALS – a post-hoc analysis of the ROCK-ALS trial

**DOI:** 10.1101/2025.09.02.25334770

**Authors:** Andreas W Wolff, Andreas Leha, Jan C. Koch, Antonia F. Demleitner, Christoph Neuwirth, Tim Friede, Markus Weber, Paul Lingor

**Author notes:** Corresponding Author: Paul Lingor, Ismaninger Str. 22, 81675, Munich, Germany.

## Abstract

Amyotrophic lateral sclerosis (ALS) is a progressive neurodegenerative disease characterized by the spread of muscle weakness across body regions. The ROCK-ALS trial was a multicenter, randomized, double-blind, placebo-controlled phase 2 study assessing the safety, tolerability, and efficacy of the Rho kinase inhibitor fasudil as an add-on to riluzole in ALS patients. A key exploratory objective was to evaluate fasudil’s effect on the spread of muscle weakness using the Motor Unit Number Index (MUNIX), a quantitative electrophysiological biomarker of lower motor neuron integrity.

MUNIX was assessed in 10 muscles (5 on each body side) at baseline, day 26, day 90, and day 180. Correlations were assessed between baseline serum biomarkers—neurofilament light chain (NfL) and glial fibrillary acidic protein (GFAP)—and baseline clinical measures (ALSFRS-R, slow vital capacity, and MUNIX sum scores) as well as their monthly rates of change, to explore potential prognostic relationships. For the analysis of disease spreading, muscles were classified as newly affected based on MUNIX decline relative to contralateral values or prior measurements, using thresholds of ≥10%, ≥20%, or ≥30%.

118 participants were included in the intention-to-treat population, 78 had full MUNIX datasets at baseline and 67 had at least one follow-up. Baseline MUNIX sum scores correlated with subsequent ALSFRS-R decline, suggesting prognostic value. Additionally, at day 90, fasudil significantly reduced the number of newly affected muscles compared to placebo in a dose dependent manner over different thresholds.

These findings support MUNIX as a sensitive biomarker for monitoring disease spreading and demonstrate that fasudil may attenuate the progression of lower motor neuron involvement in ALS.

## Introduction

ALS is a devastating neurodegenerative disorder with a mean survival of 2-4 years after symptom onset.^1^ One of the characteristics is the insidious onset of the disease, which can occur in one of the limbs, the respiratory muscles or in the bulbar region, affecting speech and swallowing. However, as ALS is a progressive disorder, these symptoms spread from the initially affected region to other regions and result in a progressive global muscle weakness. This functional spreading is also reflected by the histological spreading of the TDP-43 pathology.^2^ A therapeutic agent that could reduce the spread of a disease would have a significant impact on the progression of the disease, as it could preserve the functions of previously unaffected muscles or limbs. Fasudil is a rho kinase inhibitor that showed promising effects on survival and motor function in the SOD1.G93A mouse model of ALS.^3,4^ The ROCK-ALS trial assessed the safety, tolerability, and efficacy of fasudil in patients with ALS. Fasudil was well tolerated and safe. Secondary endpoints analyses suggested that fasudil attenuates the decline of MUNIX in patients with ALS compared with placebo. Here, we present results on the effect of fasudil on spreading of muscle weakness quantified by the motor unit number index (MUNIX) assessment.

## Methods

### Study Design and Participants

ROCK-ALS was a phase 2, randomised, double-blind, placebo-controlled clinical trial evaluating two different doses of fasudil as an add-on therapy to riluzole in patients with Amyotrophic lateral sclerosis (ALS). The trial was conducted at 19 centres across Germany, France, and Switzerland. The study protocol was approved by the relevant ethics committees and regulatory authorities in each participating country. Lead approval in Germany was obtained from the Ethics Committee of the University of Göttingen (approval number 31/5/18). The trial was registered at ClinicalTrials.gov (NCT03792490) and Eudra-CT (2017-003676-31). The study is now completed and the main results have been reported previously.^5^

Adults aged 18 to 80 years with a diagnosis of probable, laboratory-supported probable, or definite ALS according to the revised El Escorial criteria, and a disease duration of more than 6 months but less than 24 months, were eligible. Co-treatment with oral riluzole (50 mg twice daily) was mandatory. Inclusion required a predicted slow vital capacity (SVC) of >65%. Key exclusion criteria included tracheostomy or assisted ventilation in the preceding 3 months, gastrostomy, known arterial hypotension (<90/60 mm Hg), and personal or family history of intracranial bleeding, intracerebral aneurysm, or Moyamoya disease. All participants provided written informed consent prior to enrolment.

### Intervention

Fasudil hydrochloride hydrate (15 mg/mL ampoules; Eril, Asahi Kasei Pharma, Tokyo, Japan) was administered intravenously after dilution in 100 mL of 0.9% sodium chloride. Participants received either 30 mg fasudil (2 × 1 mL fasudil), 15 mg fasudil (1 mL fasudil + 1 mL NaCl), or placebo (2 × 1 mL NaCl). The investigational product was infused twice daily over 45 minutes using a CE-certified infusion pump, with a 6–8-hour interval between doses for 20 consecutive working days, excluding weekends and holidays. The high dose fasudil group received a cumulative dose of 60 mg/day for 20 days, the maximum licensed cumulative dose.

### Outcomes

Secondary efficacy endpoints of the ROCK-ALS trial included changes from baseline to days 26, 90, and 180 in the following assessments: ALS Functional Rating Scale–Revised (ALSFRS-R), ALS Assessment Questionnaire (ALSAQ-5), Edinburgh Cognitive and Behavioral ALS Screen (ECAS), Motor Unit Number Index (MUNIX), and predicted slow vital capacity (sVC).

MUNIX is a non-invasive and rapid, electrophysiological method used to estimate the number of motor units in individual muscles. It can be applied to any proximal or distal muscle from which a compound muscle action potential (CMAP) can be elicited through supramaximal nerve stimulation. The procedure consists of three steps: (1) recording the CMAP using standard motor nerve conduction techniques; (2) recording surface interference pattern (SIP) electromyography signals during varying levels of voluntary isometric contraction; and (3) calculating the MUNIX value through regression analysis of CMAP and SIP signal parameters. The method yields an “ideal case motor unit count” (ICMUC), from which MUNIX and the Motor Unit Size Index (MUSIX) are derived. Measurements are performed under standardized filter settings, and signal quality is manually reviewed to exclude artifacts or tremors. MUNIX has demonstrated good intra- and inter-rater reliability^6–9^ and is particularly suited for multicenter longitudinal studies such as ALS trials^5,7,10^, especially when combining multiple muscles into a composite “megascore” to track disease progression.

In this study, MUNIX was measured in five muscles on each side (biceps brachii, abductor pollicis brevis, abductor digiti minimi, tibialis anterior, and extensor digitorum brevis) and summarized as the MUNIX megascore 10 (MUNIX-10 score). Assessments were performed by certified raters trained in MUNIX methodology. MUNIX values were measured for each muscle at baseline and at the end of the treatment period (day 26). Follow-up MUNIX assessments were conducted at day 90 and day 180 post-baseline.

For MUNIX, the lower limit of detection (LLOD) was defined as one-fifth of the published 5% quantiles. Values below the LLOD were imputed as 50% of the LLOD. To evaluate disease spreading, all 10 muscles were assessed. At baseline, MUNIX score was defined as the mean of measurements at baseline and day 26. A muscle was classified as “affected” if it met one of the following criteria: (1) MUNIX value below the published 5% quantile; (2) ≥10%, ≥20%, or ≥30% reduction compared to the contralateral side; or (3) ≥10%, ≥20%, or ≥30% decline compared to the prior measurement. Once classified as affected, a muscle remained so for the duration of the study.

ROCK-ALS encompassed a comprehensive biomaterial collection. Serum samples were collected at baseline, at the end of the treatment period as well as on day 180 post-baseline. Serum Neurofilament-light chain (NfL) and serum glial fibrillary acidic protein (GFAP) were measured by SIMOA immunoassay on a HD-X (NEUROLOGY 2-PLEX B, Quanterix, Lexington, MA, USA).

### Statistical Analysis

Statistical analyses were performed using R Version 4.3.1. Correlations between clinical parameters (MUNIX, ALSFRS-R, sVC) and serum biomarkers (NfL and GFAP) were assessed using Spearman’s rank correlation coefficient rho. To assess the discriminative ability of individual biomarkers and composite scores (NfL/MUNIX-10 ratio) in identifying fast progressors, we performed receiver operating characteristic (ROC) curve analyses using the pROC package. The area under the curve (AUC) was calculated to quantify model performance, where an AUC of 0.5 indicates no discriminative ability and an AUC of 1.0 reflects perfect classification. We applied the Wilcoxon rank-sum test to assess whether the AUC was significantly greater than 0.5. To assess predictive performance, we performed 5-fold cross-validation, stratified by the binary outcome. For each fold, an age-, sex-, and treatment group-adjusted logistic regression model was fitted on the training set. The fitted model was applied to the held-out test set in each fold to obtain predicted probabilities. Receiver operating characteristic (ROC) curves were generated for each test fold, and the area under the ROC curve (AUC) was calculated with 95% confidence intervals (CI) using the DeLong method. The mean AUC and average 95% CIs were calculated across all folds to summarize out-of-sample model performance. Herein, the package “rsample” was used for cross-validation. For the analysis of the effect of fasudil on disease spreading assessed by MUNIX, generalized linear mixed-effects logistic regression models (GLMM) were used to examine the association between treatment group and the binary outcome “newly affected muscle”. Models accounted for the repeated measures per subject by including a random intercept for each subject to capture within-subject correlation. Odds ratios (ORs) and 95% CIs for fixed effects were obtained by exponentiating the model coefficients. Statistical significance was determined using Wald z-tests. Here, the “lme4” and “broom.mixed” packages were used. If not stated otherwise, correction for family-wise error rate was performed using the Bonferroni-Holm method. A two-sided significance level of 0.05 was used throughout.

## Results

### Baseline correlation of serum and clinical biomarkers

As previously reported^5^, a total of 118 participants received the investigational medicinal product and were therefore selected for the analysis set of the ROCK-ALS study, which is also used for the present analysis. In 78 participants (66%) all 10 muscles were assessed by MUNIX at baseline. Additionally, for 106 participants (90%) serum NfL and GFAP values were available at baseline. As shown in figure 1, correlation analysis at baseline found an intercorrelation of serum biomarkers NfL and GFAP (Spearman’s rho = 0.38; *p*_*adj*_ *<* 0.001), as well as clinical parameters sVC and ALSFRS-R (rho = 0.51; *p*_*adj*_ *<* 0.001). The MUNIX-10 sum score at baseline did not correlate with serum biomarkers (NfL or GFAP) or the ALSFRS-R score at baseline, however it showed trend towards a positive correlation with sVC (rho = 0.28; *p*_*raw*_ = 0.01, *p*_*adj*_ = 0.11)

**Figure 1.**
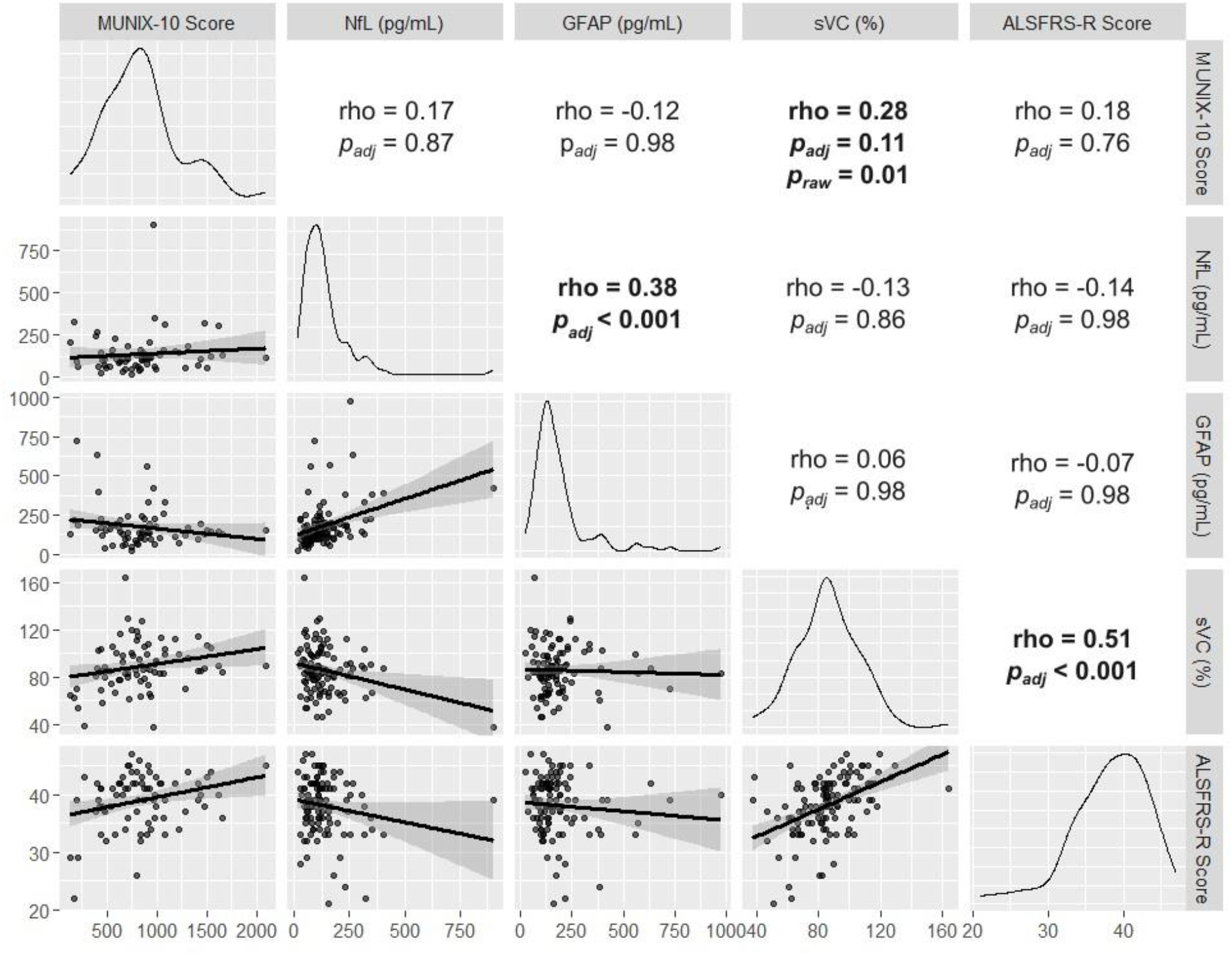
Pairwise correlations among baseline biomarkers and clinical measures in ALS. Scatter plots, density plots, and Spearman correlation coefficients are shown for MUNIX-10 score, NfL (pg/mL), GFAP (pg/mL), sVC (%), and ALSFRS-R score. Statistically significant correlations are indicated in bold. P-value adjustment was performed by Bonferroni-Holm method.

### Change in MUNIX over time

Of all 78 patients with complete MUNIX measurements (including all 10 muscles), 67 participants (86%) had at least one complete follow-up assessment, either after 3 months or after 6 months. MUNIX-10 sum score declined over time (baseline: 831 +/−376 (mean score +/−standard deviation); after 3 months: 705 +/−349; after 6 months: 637 +/−334). The individual change in the MUNIX-10 sum score was calculated by subtracting the baseline value from the last data point, divided by the number of months between the baseline and the last data point. Baseline ALSFRS-R, baseline sVC and GFAP did not correlate with change in MUNIX-10 score. However, baseline NfL correlated with change in MUNIX-10 sum score (rho= −0.35, *p*_*adj*_ = 0.02, Figure 2). We found that higher baseline MUNIX-10 score correlated with more pronounced loss in MUNIX-10 sum score (rho = −0.36; *p*_*adj*_ = 0.01), while higher baseline ALSFRS-R correlated with slower decline of ALSFRS-R (rho = 0.30; *p*_*adj*_ < 0.001). The proportional monthly change rate relative to baseline MUNIX-10 score showed no significant correlation with the baseline MUNIX-10 score (rho = 0.80; *p*_*adj*_ = 1), indicating that changes over time were not associated with initial values. In contrast, the proportional change from baseline for the ALSFRS-R score was significantly correlated with the baseline ALSFRS-R score (rho = 0.42; *p*_*adj*_ < 0.001), suggesting that the rate of change was related to patients’ baseline functional status.

**Figure 2.**
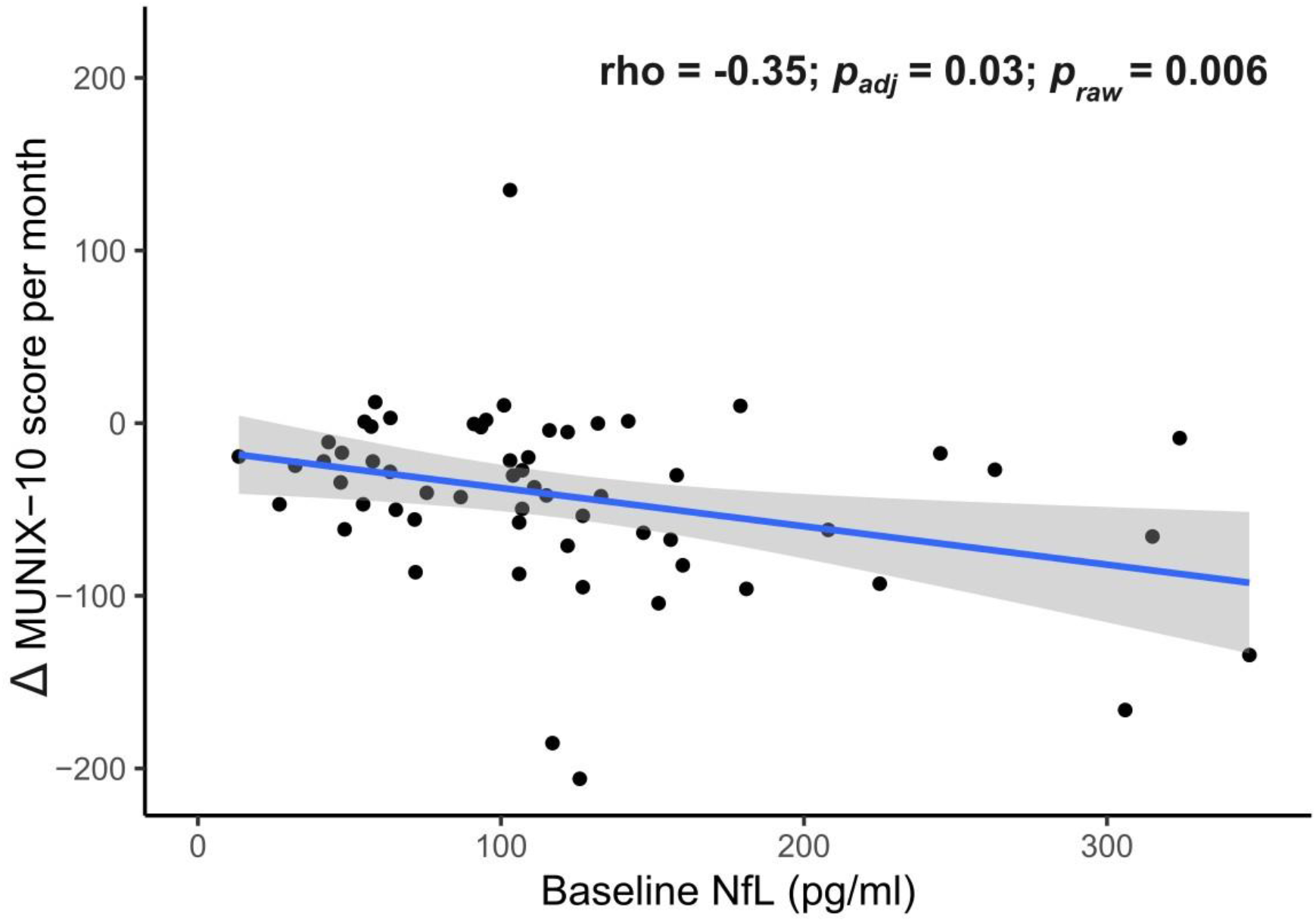
Association between baseline NfL and MUNIX-10 scores change per month Blue line indicates linear regression fit; shaded areas represent 95% confidence intervals. Corresponding Spearman correlation coefficient (rho) with p-values (Bonferroni-Holm adjusted and unadjusted) is indicated.

### Prognostic value of MUNIX

Next, we aimed to assess the prognostic potential of serum biomarkers and MUNIX. We deemed change in slow vital capacity and ALSFRS-R clinically relevant outcome parameters. Baseline GFAP did not significantly correlate with the change in sVC per month (rho = 0.04, *p*_*adj*_ = 0.69) or ALSFRS-R change per month (rho = 0.11, *p*_*adj*_ = 0.66). Baseline NfL, however, correlated negatively with change in slow vital capacity per month and ALSFRS-R change per month, respectively (Figures 3A and 3B).

**Figure 3.**
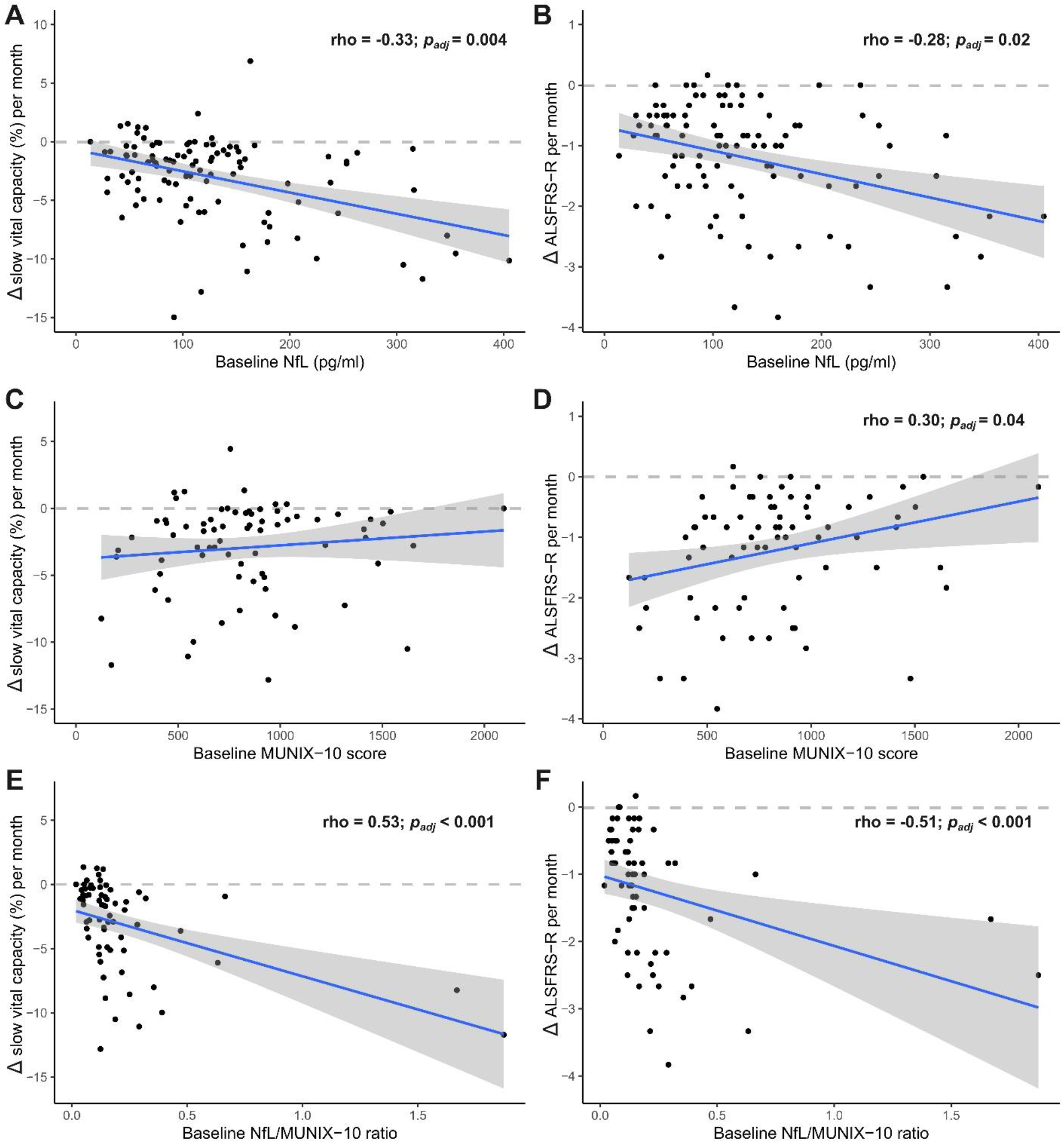
Correlations between baseline biomarkers and clinical decline in ALS. (A, B) Higher baseline NfL levels (pg/mL) are associated with greater monthly decline in slow vital capacity (Δ sVC %/month) and ALSFRS-R (Δ ALSFRS-R/month), respectively. (C, D) Higher baseline MUNIX-10 scores are associated with slower decline in ALSFRS-R (D), but show no significant association with Δ SVC (C). The most pronounced correlations were found for baseline NfL/MUNIX-10 ratio with Δ SVC (E) and Δ ALSFRS-R (F). Each panel shows individual data points, linear regression lines (blue), 95% confidence intervals (gray shading), and corresponding Spearman correlation coefficients (rho) with p-values adjusted for multiplicity by Bonferroni-Holm.

The baseline MUNIX-10 sum score did not correlate with change in slow vital capacity per month, but with ALSFRS-R change per month (Figures 3C and 3D), even to a greater extent than NfL. To evaluate a potential additive predictive value of NfL and MUNIX-10, the ratio of NfL and MUNIX-10 score was calculated. The NfL/MUNIX-10 ratio correlated numerically better with the monthly change of ALSFRS-R and monthly change of sVC compared to NfL and MUNIX-10 score alone (Figure 3E and 3F).

### Additive prognostic value of MUNIX and NfL

We then aimed to further strengthen evidence on a potential additive value of NfL and MUNIX-10 to predict progression. In an age-, sex-, and fasudil-treatment-adjusted multivariable linear regression model both NfL (β (95%-CI) = −0.01 (−0.01 to −0.00), p < 0.001) and MUNIX-10 score (β (95%-CI) = 0.00 (0.00 to 0.00), p = 0.003) were identified as independent significant predictors for ALSFRS-R change. This effect was also identified on the sVC change (NfL (β (95%-CI) = −0.02 (−0.03 to −0.01), p < 0.001) and MUNIX-10 score (β (95%-CI) = 0.00 (0.00 to 0.00), p = 0.045), respectively). We then evaluated whether baseline NfL/MUNIX-10 ratio, MUNIX-10 score or NfL values could discriminate between fast and slow progressors based on their rate of functional decline in ALSFRS-R or sVC, as assessed by receiver operating characteristic (ROC). Individuals with a monthly decline rate equal or above the group median (−1 points per month in ALSFRS-R and −1.83% loss of sVC per month) were classified as fast progressors. For ALSFRS-R, the NfL/MUNIX-10 ratio numerically outperformed NfL or MUNIX-10 score individually. Here, the area under the ROC curve (AUC) of the NfL/MUNIX-10 ratio was 0.79 (95% CI: 0.68–0.90, DeLong method), indicating numerically better discriminative ability compared to NfL (AUC = 0.74; 95% CI: 0.62-0.86) and MUNIX-10 (AUC = 0.69; 95% CI: 0.57-0.82), alone. For sVC, the AUC of NfL/MUNIX-10 ratio was 0.74 (95% CI: 0.62–0.87) compared to NfL (AUC = 0.76; 95% CI: 0.64-0.87) and MUNIX-10 (AUC = 0.65; 95% CI: 0.51-0.78), alone (Fig 4). The discriminative power of the model was validated using stratified 5-fold cross-validation. The mean cross-validated AUC of the NfL/MUNIX-10 ratio to identify fast progressors in ALSFRS-R was AUC 0.71 (95% CI: 0.41-0.98; p = 0.03, Wilcoxon signed rank exact test for greater than 0.5). For sVC, the mean discriminative power across all folds of the NfL/MUNIX-10 ratio was AUC 0.65 (95% CI: 0.32-0.96; p = 0.03).

**Figure 4:**
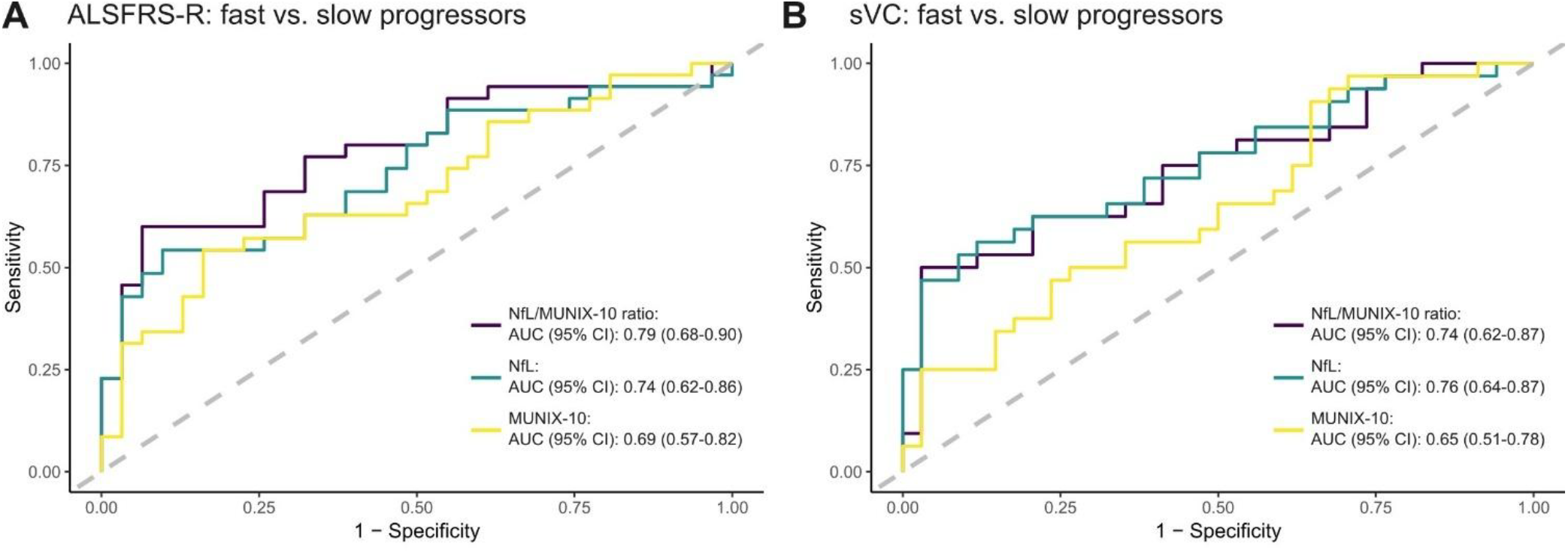
Receiver operating characteristic (ROC) curves comparing the predictive performance of age-, sex-, and fasudil-treatment-adjusted models for distinguishing fast vs. slow progressors based on the decline of ALSFRS-R (A) and sVC (B). ROC curves, as well as AUC and 95% CI are displayed for NfL/MUNIX-10 ratio (purple), NfL only (teal), and MUNIX-10 only (yellow).

### Spreading analysis

At baseline, any MUNIX data were available for 98 of 118 enrolled participants. Any follow-up data were available for 89 participants on day 90 and for 70 participants on day 180. This corresponds to 836 individual muscles with at least one follow-up measurement: 309 muscles in the placebo group, 253 muscles in the 15 mg fasudil group, and 274 muscles in the 30 mg fasudil group.

To assess disease progression, three relative change thresholds (≥10%, ≥20%, or ≥30%) were applied to identify affected muscles. These were selected based on available literature on the inter- and intra-rater variance of MUNIX. A muscle was classified as “affected” if its MUNIX value fell below these thresholds relative to either its contralateral counterpart or its previous measurement, thereby exceeding the expected re-measurement variance. Generalized linear mixed-effects logistic regression model were used to analyze the probability of newly affected muscles per timepoint between fasudil treatment compared to placebo, while accounting for the repeated measures per subject by including a random intercept for each subject to capture within-subject correlation.

Across all thresholds and treatment arms, the number of newly affected muscles increased progressively from baseline to day 90, and again from day 90 to day 180 (Figure 5).

**Figure 5.**
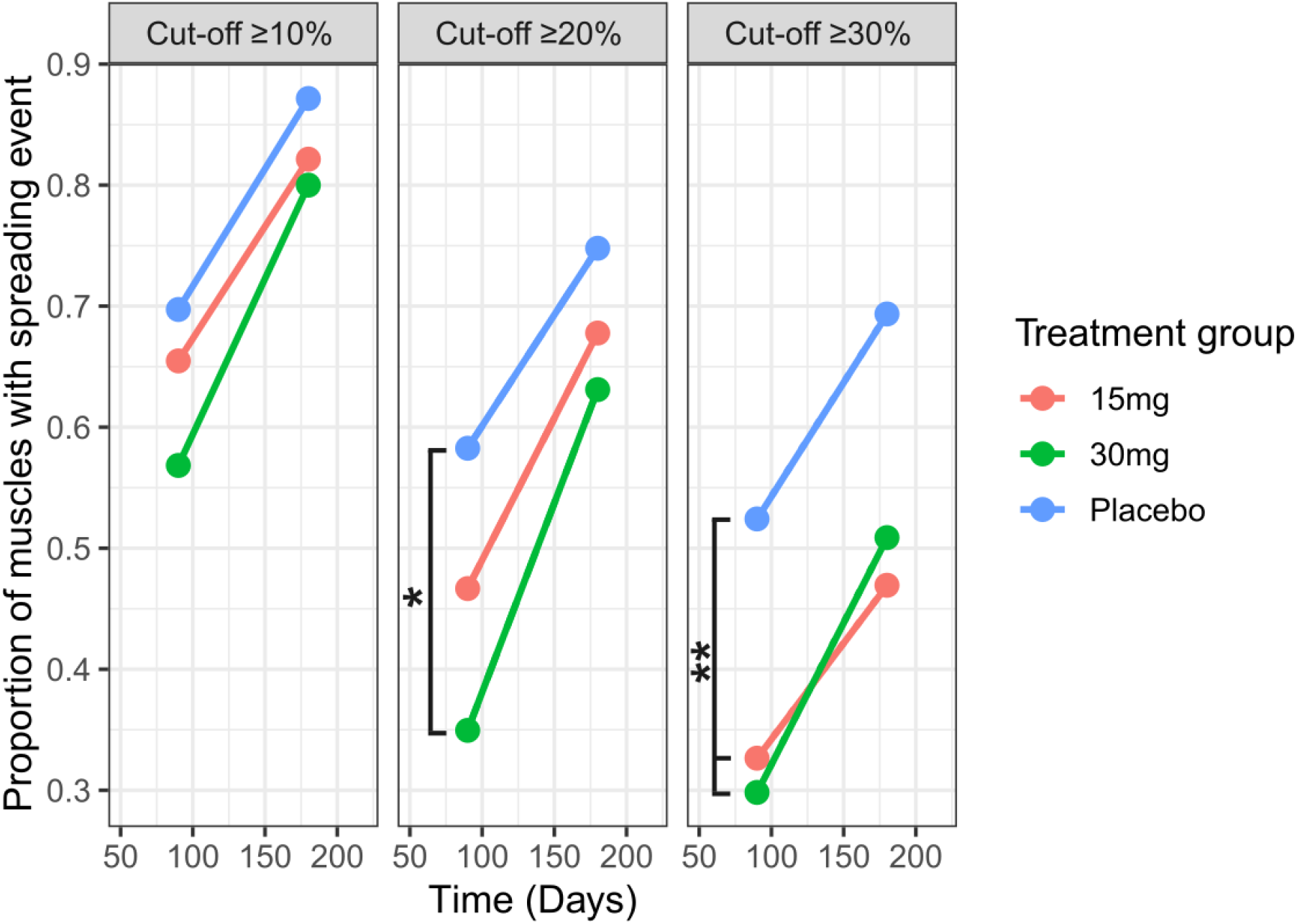
Proportion of muscles with new spreading events over time by treatment group and MUNIX cut-off. Proportion of muscles with newly defined spreading events between baseline and day 90, as well as between day 90 and day 180. Treatment groups: placebo (blue), fasudil 15 mg (red), and fasudil 30 mg (green). Cut-off thresholds are given at ≥10%, ≥20%, ≥30%. Generalized linear mixed-effects logistic regression model (GLMM) including a random intercept for each subject to capture within-subject correlation were used to assess the effect of treatment group on newly affected muscles. Odds ratios (ORs) and 95% confidence intervals (CIs) for fixed effects were obtained. Correction for family-wise error rate was performed using the Benjamini-Hochberg method. *30mg vs. placebo: OR 0.36 (95% CI: 0.17 to 0.74; *p*_*adj*_ = 0.035, *p*_*raw*_ = 0.006) ** 30mg vs. placebo: OR 0.36 (95% CI: 0.17 to 0.74; *p*_*adj*_ = 0.035, *p*_*raw*_ = 0.006); 15mg vs. placebo OR 0.42 (95% CI: 0.20 to 0.89; *p*_*adj*_ = 0.09, *p*_*raw*_ = 0.023).

At the 30% threshold, baseline unaffected muscle counts were 124 (42.5%) in the placebo group, 98 (43.9%) in the fasudil 15 mg group, and 114 (42.5%) in the fasudil 30 mg group. At day 90, the odds of newly affected muscles was significantly reduced in the fasudil groups compared to placebo, corresponding to an odds ratio (OR) of 0.36 (95% CI: 0.17 to 0.74; p_adj_ = 0.029, p_raw_ = 0.006) for the 30mg and OR 0.42 (95% CI: 0.20 to 0.89; p_adj_ = 0.11, p_raw_ = 0.023) for the 15mg group, respectively. This effect was negated on day 180.

At the 20% threshold, the number of unaffected muscles at baseline was 115 (39.3%) in the placebo group, 90 (40.4%) in the fasudil 15 mg group, and 103 (38.4%) in the fasudil 30 mg group. At day 90, the odds of newly affected muscles were significantly reduced in the 30 mg fasudil group compared to placebo. For the 30mg group the OR was 0.36 (95% CI: 0.17 to 0.74; p_adj_ = 0.034, p_raw_ = 0.006) and for the 15 mg the OR was 0.61 (95% CI: 0.29 to 1.28; p_raw_ = 0.19, p_adj_ = 1). Again, this effect was not observed on day 180.

For the 10% threshold, a similar trend was observed: fasudil treatment groups exhibited fewer newly affected muscles on day 90, but the differences did not reach statistical significance.

## Discussion

The ROCK-ALS trial was the first placebo-controlled, double-blind, randomized phase II trial to evaluate the ROCK inhibitor fasudil in patients with amyotrophic lateral sclerosis (ALS), with a primary focus on safety and tolerability.^5^ The trial met its primary endpoint, safety and tolerability, demonstrating that fasudil was well tolerated, with no serious adverse events attributed to the drug. Its secondary endpoints included ALSFRS-R, survival, cognitive and behavioral assessment using the Edinburgh Cognitive Assessment Screen (ECAS), slow vital capacity (sVC) as well as the MUNIX megascore 10. Fasudil showed a significant dose-dependent effect, attenuating MUNIX decline at day 45 and 90. Exploratory analyses showed a sex-specific effect of fasudil on the attenuation of sVC decline. These findings, although limited by the short treatment duration and lack of multiplicity adjustment, suggest disease-modifying effects of fasudil that warrant further investigation.

In the present analysis, we aimed to further explore the potential of MUNIX as an outcome measure in ALS clinical trials investigating disease-modifying therapies. The longitudinal utility of MUNIX for monitoring disease progression has been demonstrated in both limb- and bulbar-onset ALS.^12^ The ALSFRS-R, a widely used clinical endpoint, represents a functional composite score influenced by both upper and lower motor neuron involvement and additional factors (e.g., non-invasive ventilation, PEG feeding, symptomatic interventions such as for sialorrhoea). In contrast, MUNIX reflects lower motor neuron loss more directly and is less susceptible to such confounding variables, supporting its use as a complementary objective biomarker—particularly in phase II trials. In our data, baseline ALSFRS-R and slow vital capacity were strongly correlated, while MUNIX-10 correlated only in trend with slow vital capacity, consistent with recent cross-sectional findings in ALS.^13^ Evidence on the correlation between ALSFRS-R and MUNIX-10 is inconsistent, with studies reporting both positive correlations^13–16^ and no correlation^17^, likely due to differences in patient populations and muscles assessed. In our study, we observed a trend towards a positive correlation between ALSFRS-R and MUNIX-10 scores, although they did not reach statistical significance. This further supports the concept that MUNIX provides complementary, independent information on disease severity in ALS.

The decline of ALSFRS-R and MUNIX scores over time has been well documented.^12,13^ In ROCK-ALS, ALSFRS-R showed a mean reduction of ~8% at 3 months and ~16% at 6 months.^5^ In our extended analysis, the MUNIX-10 sum score declined by ~15% in 3 months and ~23% in 6 months, aligning with prior evidence that MUNIX detects greater relative decline than ALSFRS-R and may be more sensitive to subtle changes over short intervals. Moreover, we found that the baseline ALSFRS-R correlated with relative decline of the individual baseline ALSFRS-R, whereas no such association was observed for MUNIX-10. This confirms that ALSFRS-R progression is influenced by baseline functional status, which has already been demonstrated in a meta-analysis including over 2,000 patients.^18^ In contrast, relative MUNIX decline appeared more stable and independent of individual baseline measurements, reinforcing its potential as an objective marker of lower motor neuron loss.

Additionally, baseline MUNIX-10 scores correlated with subsequent ALSFRS-R decline, suggesting that early MUNIX assessment may help predict disease progression. This supports the notion that lower motor neuron loss can precede measurable functional decline, particularly in early stages when subtle symptoms may not yet manifest in reduced ALSFRS-R. Patients with low baseline MUNIX values may thus be more likely to experience faster clinical deterioration. MUNIX has also been shown to predict 1-year survival with good sensitivity and moderate specificity.^13^ To our knowledge, this is the first study demonstrating that baseline MUNIX scores can predict subsequent ALSFRS-R decline. The ratio of NfL and MUNIX outperformed the individual biomarkers alone in identifying fast progressors as assessed by ALSFRS-R. This strengthens the argument that MUNIX, as marker of the lower motor neuron, can provide complementary information about disease severity and trajectory in addition to NfL.

ALS progression involves not only symptom worsening but also the spread of pathology to new body regions.^19^ Previously approved ALS treatments, including riluzole and edaravone, have shown modest effects on functional decline and survival.^1^ Fasudil was shown to attenuate MUNIX decline after 3 months in a dose-dependent fashion. However, its potential effect on disease spread has not yet been evaluated in clinical trials. Given MUNIX’s sensitivity to detect subclinical involvement of new muscle groups, we examined fasudil’s ability to limit disease spread. Muscles were classified as “not affected” if MUNIX values exceeded established thresholds^6^ or if there were no significant side-to-side differences. We then tracked the emergence of newly affected muscles after 3 and 6 months, applying three thresholds based on inter- and intra-rater reproducibility data. ^6,7^ Fasudil reduced the number of newly affected muscles by 3 months in a dose-dependent manner, with effects replicated across two threshold definitions. This effect was attenuated by 6 months, suggesting a short-term benefit in slowing disease spread. Limiting the spread of lower motor neuron degeneration could offer significant therapeutic benefits by preserving unaffected regions.

Our study has several limitations. Most importantly, fasudil had to be administered intravenously due to current labeling, restricting treatment duration and dosage, which may have limited efficacy. However, fasudil has good oral bioavailability, and an oral formulation may facilitate chronic administration in future trials. ^20^ The study was not powered for efficacy endpoints, and no long-term follow-up was conducted. While MUNIX shows promise as a progression biomarker, it requires standardization and trained personnel to mitigate variability. We addressed this by providing detailed manuals, on-site training, and test–retest reliability checks to ensure consistency across centers.

## Conclusion

This study demonstrates the relevance of MUNIX as a biomarker of disease progression and a potential outcome parameter in ALS clinical trials. Importantly, our findings reinforce that fasudil may exert disease-modifying effects on lower motor neurons. Furthermore, we provide the first evidence that fasudil can reduce disease spread to previously unaffected muscle regions, highlighting its potential therapeutic impact.

## Data availability

De-identified individual participant data will be available 12 months after publication via a restricted-access, online data repository accessible at https://doi.org/10.25625/HOZIRN. These data will include demographics, vital signs, ALSFRS-R, SVC, ALSAQ-5, MUNIX, ECAS (total scores, each), and others, including a data dictionary. The study protocol is available as an open access publication. Data will be available for researchers or investigators whose use has been reviewed and approved by the ROCK-ALS data access committee to be used in scientific analyses of the individual data or for merging with other data in meta-analyses. Applicants must sign a data access agreement. Requests should be addressed via email to the corresponding author.

